# Necessity of COVID-19 Vaccination in Persons who have Already had COVID-19

**DOI:** 10.1101/2021.06.01.21258176

**Authors:** Nabin K. Shrestha, Patrick C. Burke, Amy S. Nowacki, Paul Terpeluk, Steven M. Gordon

## Abstract

**Background:** The purpose of this study was to evaluate the necessity of COVID-19 vaccination in persons with prior COVID-19.

**Methods:** Employees of Cleveland Clinic working in Ohio on Dec 16, 2020, the day COVID-19 vaccination was started, were included. Anyone who tested positive for COVID-19 at least once before the study start date was considered previously infected. One was considered vaccinated 14 days after receipt of the second dose of a COVID-19 mRNA vaccine. The cumulative incidence of COVID-19, symptomatic COVID-19, and hospitalizations for COVID-19, were examined over the next 10.5 months.

**Results:** Among the 52238 employees, 4718 (9%) had prior COVID-19 at the start of the study, and 35113 (67%) had received at least two doses of the vaccine by the end of the study. Of the 4284 COVID-19 infections during the study, 3476 (81.1%) occurred in persons who were unvaccinated, and 4263 (99.5%) occurred among those without prior COVID-19. In Cox proportional hazards regression, both prior COVID-19 and vaccination were independently associated with significantly lower risk of COVID-19. Vaccination was associated with lower risk of COVID-19 among those without prior COVID-19 (HR 0.24, 95% CI 0.22–0.26) but not among those with prior COVID-19 (HR 0.86, 95% CI 0.33–2.29).

**Conclusions:** Both previous infection and vaccination provide substantial protection against COVID-19. Vaccination reduces risk of COVID-19 among those without prior COVID-19 but not among those with prior COVID-19, at least not within one year following infection.

**Summary:** Cumulative incidence of COVID-19 over 10.5 months, including the Delta phase, was examined among 52238 employees at Cleveland Clinic. Vaccination was associated with significantly lower risk of COVID-19 among those without prior COVID-19 but not among those with prior COVID-19.

## INTRODUCTION

We have previously reported that individuals who have previously had coronavirus disease of 2019 (COVID-19), caused by Severe Acute Respiratory Syndrome (SARS) – associated Coronavirus-2 (SARS-CoV-2), are unlikely to benefit from vaccination against COVID-19 [1]. Since then, a new variant, the Delta variant has spread worldwide, and questions have been raised about whether our findings continue to hold true at a time when the Delta variant is predominant, and whether protection from prior infection lasts longer than a few months. An unnecessarily restrictive definition of prior infection in our previous study also misclassified persons infected in the recent past as previously uninfected. The purpose of this study was to re-examine the question about whether individuals with prior COVID-19 benefit from getting vaccinated, over a longer follow-up period and during the Delta phase of the pandemic.

## METHODS

### Study design

This was a retrospective cohort study conducted at the Cleveland Clinic Health System in Ohio, USA. The study was approved by the Cleveland Clinic Institutional Review Board as exempt research (IRB no. 21-985). A waiver of informed consent and waiver of HIPAA authorization were approved to allow access to de-identified health information by the research team.

### Setting

The arrival of the COVID-19 pandemic brought unprecedented stress to the Cleveland Clinic Health System, as it did to all other healthcare institutions, and identification and management of infected employees took on great importance and urgency. PCR testing for SARS-CoV-2 at Cleveland Clinic began on March 12, 2020, and a streamlined process dedicated to the testing of health care personnel (HCP) was begun shortly thereafter. All employees with a positive SARS-CoV-2 test were interviewed and symptoms monitored remotely by Occupational Health while the employees were not hospitalized.Voluntary vaccination for COVID-19 began at Cleveland Clinic on December 16, 2020. Most employees received an mRNA vaccine, either the Pfizer-BioNTech vaccine or the Moderna vaccine. All employees were scheduled to receive their second vaccine dose 28 days after the first one, regardless of which vaccine was given. Documentation of COVID-19 vaccination and of any SARS-CoV-2 test in the Occupational Health database, made the employee cohort a suitable cohort to examine the protective effect of vaccination and prior COVID-19 against future infection.

### Participants

All employees of the Cleveland Clinic Health System, working in Ohio were screened for inclusion in the study. Those in employment on December 16, 2020, were included.

### Variables

COVID-19 was defined as a positive nucleic acid amplification test (NAAT) for the SARS-CoV-2 virus. The date of infection was taken to be the date of the first positive test for that episode of illness. A person who chose to receive the vaccine was considered vaccinated 14 days after receipt of the second dose of the vaccine (which would have been 42 days after receipt of the first dose of the vaccine for most subjects). Any person who tested positive for SARS-CoV-2 before the vaccine rollout date (December 16, 2020), was considered to have had prior COVID-19. Other covariates collected were age, aggregated job title (to maintain anonymity for rare job titles), and job location. The aggregated job title could be one of the following: professional staff, nursing, advanced practice practitioner, resident/fellow, research, pharmacy, administration, clinical support, or administrative support. The job location variable could be one of the following: Cleveland Clinic Main Campus, regional hospital (within Ohio), ambulatory center, administrative center, or remote location. A job type categorization into patient-facing or non-patient facing was done, and subjects assigned to one or the other based on aggregated job title and job location. Protected health information identifiers were not included in the extracted data, and institutional data governance rules related to employee data limited our ability to supplement our dataset with additional clinical variables.

### Outcome

The primary study outcome was time to COVID-19, the latter defined as a positive nucleic acid amplification test for SARS-CoV-2 on or after December 16, 2020. Time to COVID-19 was calculated as number of days from December 16, 2020 (vaccine rollout date, and study start date) to a subsequent positive SARS-CoV-2 test. For those with prior COVID-19, positive tests within 90 days of the first positive test were considered part of the initial episode of illness. Employees that had not developed COVID-19 were censored at the end of the study follow-up period (October 31, 2021). Those who received the Johnson & Johnson vaccine (319 subjects) without having had COVID-19 were censored on the date of receipt of the vaccine, and those whose employment was terminated during the study period before they had COVID-19 (6441 subjects) were censored on the date of termination of employment. The health system never had a requirement for systematic asymptomatic employee test screening. Most of the positive tests, therefore, would have been tests done to evaluate suspicious symptoms or, since June 21, 2021, as part of quarantine and return to work testing of employees exposed to patients with COVID-19, to remain in compliance with the Occupational Safety and Health Administration (OSHA) Healthcare Emergency Temporary Standard (final rule June 21, 2021). A small proportion would have been tests done as part of pre-operative or pre-procedural screening.

Secondary outcomes were time to symptomatic COVID-19 and time to hospitalization for COVID-19. Documentation of at least one symptom during remote home monitoring of employees, from among fever (>100.4 F), cough, shortness of breath, worsening appetite, vomiting, diarrhea, weakness, or a peripheral oxygen saturation (SpO2) < 95%, within 7 days of a positive NAAT for SARS-CoV-2, was considered symptomatic COVID-19. Time to symptomatic COVID-19 was defined as number of days from the study start date to the first positive SARS-CoV-2 test of an episode of symptomatic COVID-19. Hospitalization within 3 days before or 14 days after any positive NAAT for SARS-CoV-2, and associated occurrence of at least one symptom of COVID-19, was considered hospitalization for COVID-19. Time to hospitalization for COVID-19 was defined as number of days from the study start date to the first positive SARS-CoV-2 test of an episode of hospitalization for COVID-19. Censoring for receipt of the Johnson & Johnson vaccine and for termination of employment during the study period were done in the same manner as for the primary analysis.

### Statistical analysis

A Simon-Makuch hazard plot [2] was created to compare the cumulative incidence of COVID-19 among subjects with prior COVID-19 who were vaccinated, with those of subjects with prior COVID-19 who remained unvaccinated, subjects without prior COVID-19 who were vaccinated, and subjects without prior COVID-19 who remained unvaccinated. Vaccination was treated as a time-dependent covariate whose value changed from “unvaccinated” to “vaccinated” 14 days after receipt of a second dose of either the Pfizer or Moderna vaccine (Figure 1). Curves for the unvaccinated were based on data throughout the duration of the study for those who did not receive two doses of the vaccine, and until the date the vaccination status changed from “unvaccinated” to “vaccinated” for those who received two doses of the vaccine. Curves for the vaccinated were based on data for individuals from the date their vaccination status changed to “vaccinated”, until the study end date.

**Figure 1.**
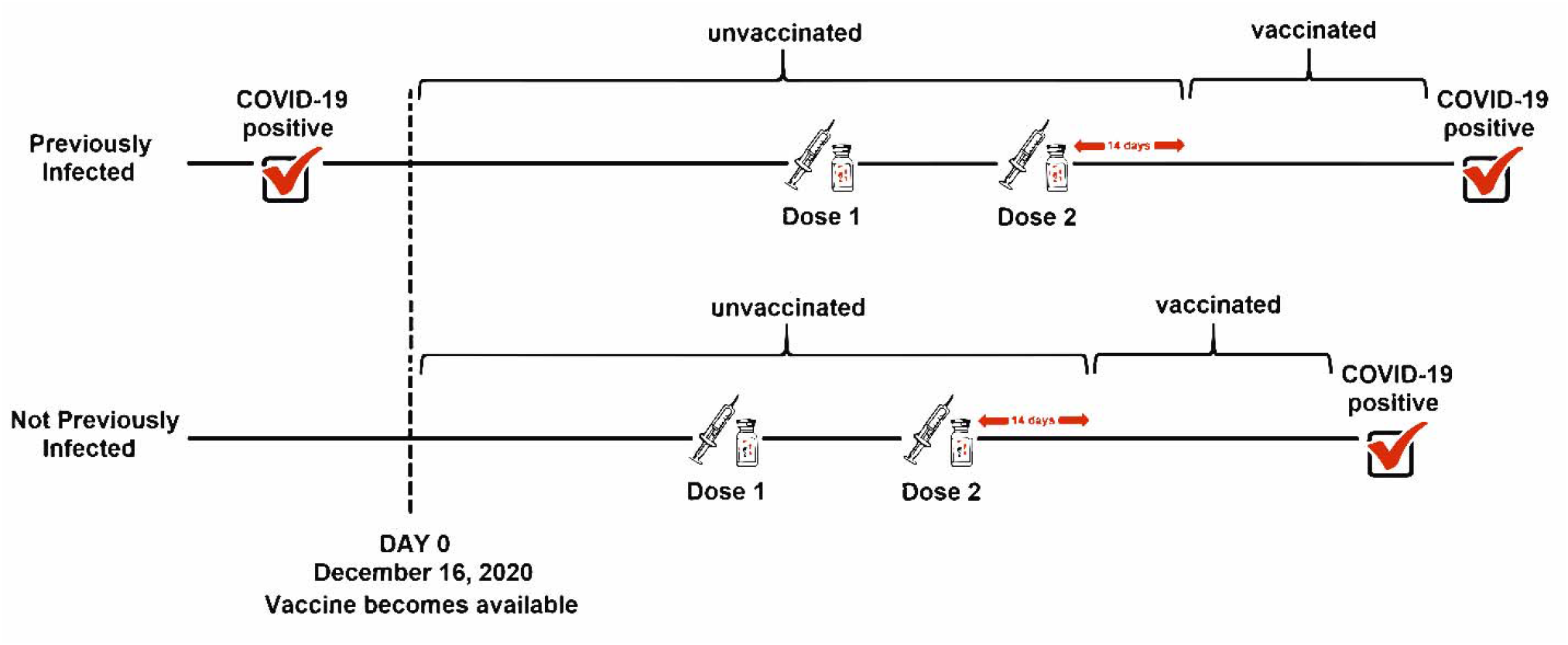
Explanation of “previously infected” analyzed as a time-independent covariate and “vaccinated” treated as a time-dependent covariate.

Simon-Makuch hazard plots were similarly constructed for time to symptomatic COVID-19 and time to hospitalization for COVID-19.

Cox proportional hazards regression models were fitted to examine associations of various variables with time to COVID-19. A multivariable model was fitted with time to COVID-19 as the outcome variable, and prior COVID-19 and vaccination (as a time-dependent covariate whose value changed on the date a subject was considered vaccinated), as explanatory variables [3]. An interaction term for prior COVID-19 and vaccination was included as a covariate. The number of covariates in the multivariable model was limited because of low numbers of events in those with prior COVID-19.

The analysis was performed by NKS and ASN using the *survival* package and R version 4.0.5 [3– 5].

## RESULTS

Of 52238 employees included in the study, 4718 (9%) had had prior COVID-19 infection, and 35113 (67%) had received at least two doses of the vaccine by the end of the study. A total of 4284 employees acquired COVID-19 during the course of the study, of which 3092 (72%) were symptomatic infections and 107 (2.5%) required hospitalization for COVID-19.

### Baseline characteristics

Those with prior COVID-19 were significantly younger (mean ± SD age; 39 ± 13 vs. 42 ± 13, p<0.001), and included a significantly higher proportion with patient-facing jobs (60% vs. 49%, p<0.001). Table 1 shows the characteristics of subjects grouped by whether or not they had prior COVID-19. A significantly lower proportion of those with prior COVID-19 (60%, 2826 subjects) were vaccinated by the end of the study compared to 68% (32228) of those not previously infected (p<0.001). Of those vaccinated, 57% received the Moderna vaccine. The epidemic in Ohio was at the peak of its third wave when vaccination was begun, and another wave peaked around 8 months later (Figure 2).

**Table 1.**
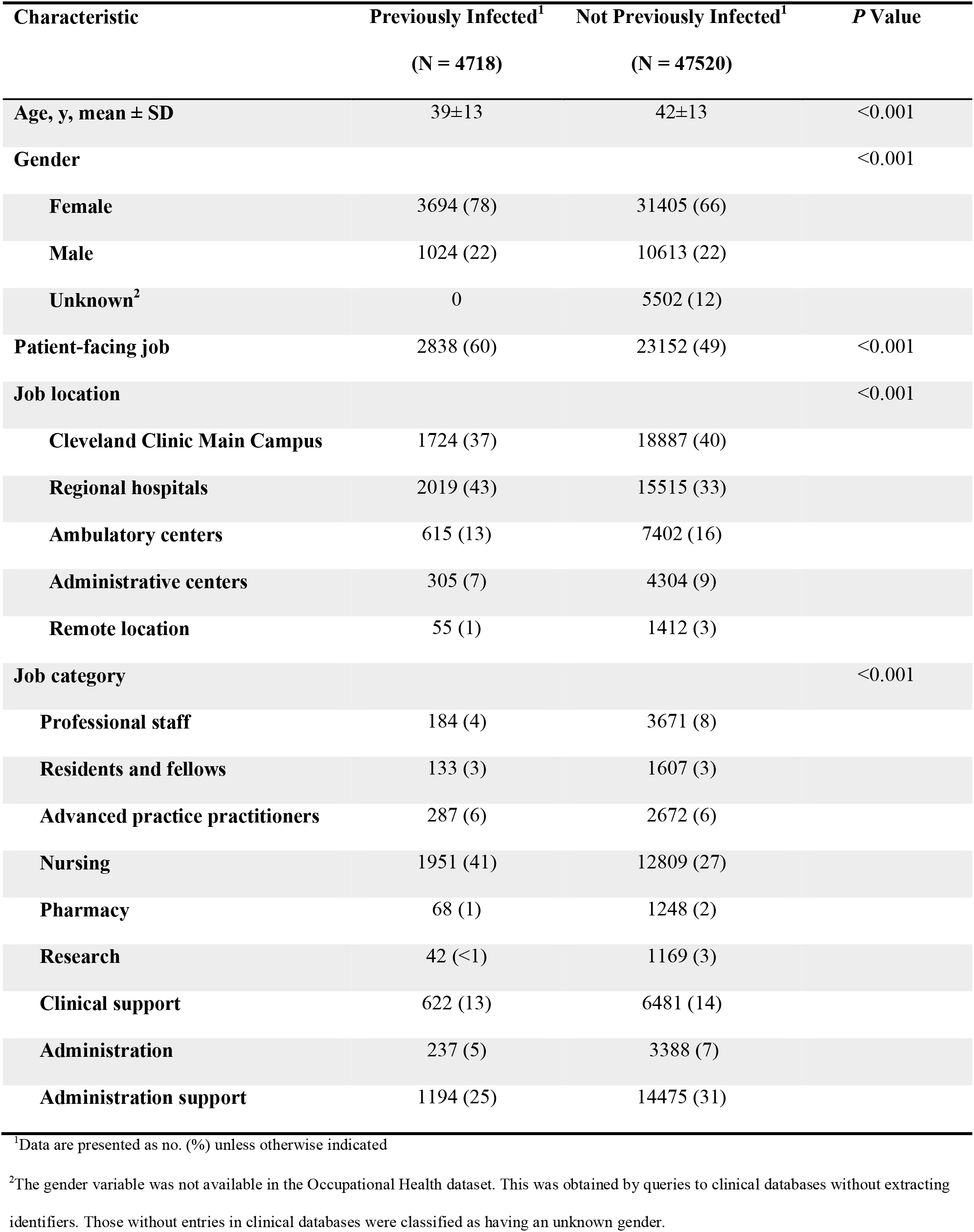
Study Subject Characteristics.

| Characteristic | Previously Infected<sup>1</sup><br>(N = 4718) | Not Previously Infected<sup>1</sup><br>(N = 47520) | P Value |
| --- | --- | --- | --- |
| Age, y, mean ± SD | 39±13 | 42±13 | <0.001 |
| Gender |  |  | <0.001 |
| Female | 3694 (78) | 31405 (66) |  |
| Male | 1024 (22) | 10613 (22) |  |
| Unknown<sup>2</sup> | 0 | 5502 (12) |  |
| Patient-facing job | 2838 (60) | 23152 (49) | <0.001 |
| Job location |  |  | <0.001 |
| Cleveland Clinic Main Campus | 1724 (37) | 18887 (40) |  |
| Regional hospitals | 2019 (43) | 15515 (33) |  |
| Ambulatory centers | 615 (13) | 7402 (16) |  |
| Administrative centers | 305 (7) | 4304 (9) |  |
| Remote location | 55 (1) | 1412 (3) |  |
| Job category |  |  | <0.001 |
| Professional staff | 184 (4) | 3671 (8) |  |
| Residents and fellows | 133 (3) | 1607 (3) |  |
| Advanced practice practitioners | 287 (6) | 2672 (6) |  |
| Nursing | 1951 (41) | 12809 (27) |  |
| Pharmacy | 68 (1) | 1248 (2) |  |
| Research | 42 (<1) | 1169 (3) |  |
| Clinical support | 622 (13) | 6481 (14) |  |
| Administration | 237 (5) | 3388 (7) |  |
| Administration support | 1194 (25) | 14475 (31) |  |

<sup>1</sup>Data are presented as no. (%) unless otherwise indicated
<sup>2</sup>The gender variable was not available in the Occupational Health dataset. This was obtained by queries to clinical databases without extracting identifiers. Those without entries in clinical databases were classified as having an unknown gender.

**Figure 2.**
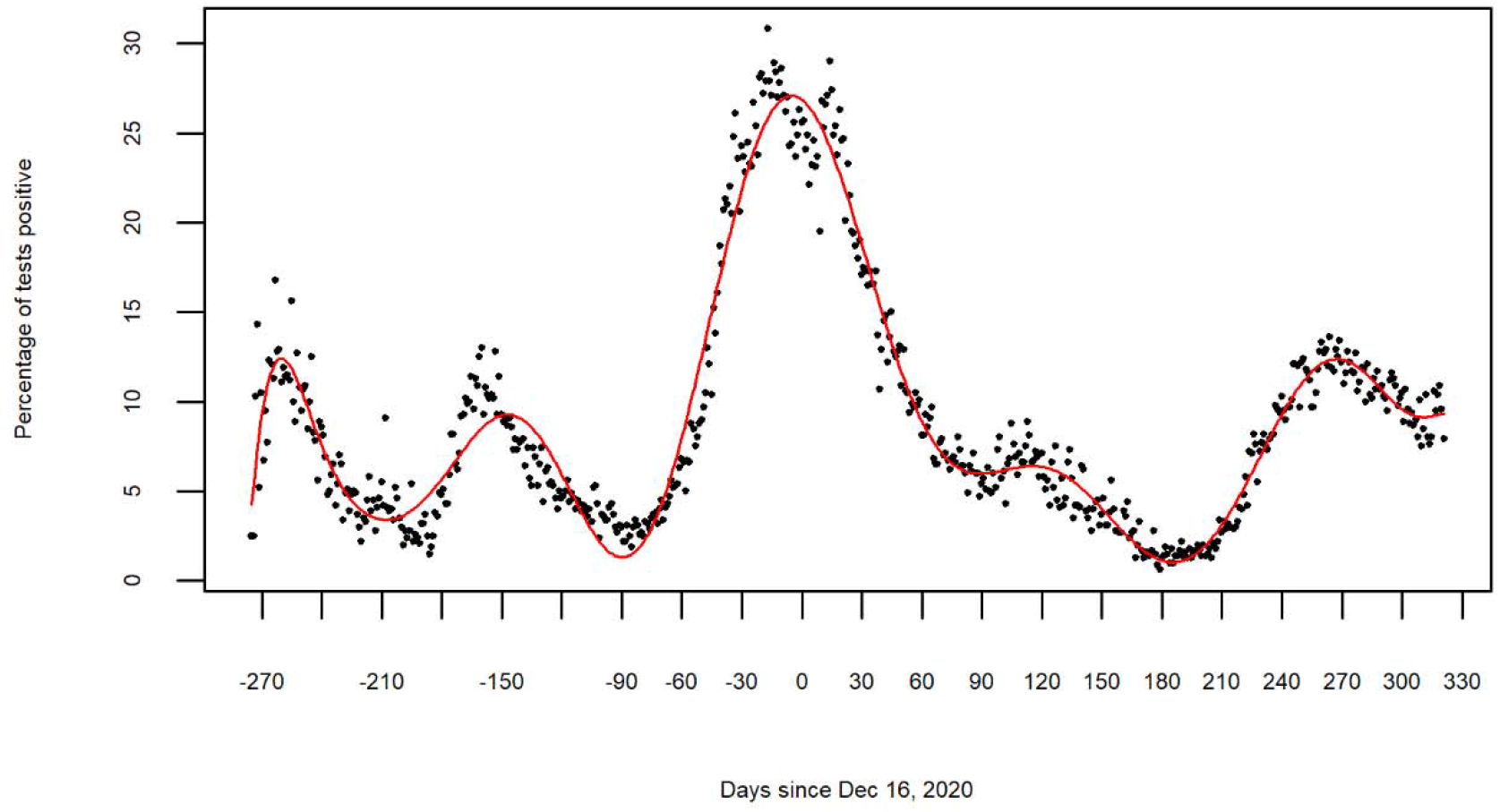
COVID-19 epidemic curve before and after the study start date. Points on the scatter plot represent the proportion of all COVID-19 PCR tests done at Cleveland Clinic that were positive on any given day. The colored line represents a fitted polynomial curve.

### Cumulative incidence of COVID-19

Figure 3 compares the cumulative incidence of COVID-19 among individuals separated by whether or not they had prior COVID-19 and whether or not they were vaccinated. The cumulative incidence of COVID-19 was highest for those without prior COVID-19 who remained unvaccinated, and was substantially lower for all others. Before the emergence of the Delta variant, the cumulative incidence of COVID-19 was negligible in those who were either previously infected or vaccinated. After the Delta variant became the predominant circulating strain, those without prior COVID-19 who remained unvaccinated continued to have the highest incidence of COVID-19. Those without prior COVID-19 who received the vaccine had a significantly lower incidence of COVID-19. Those with prior COVID-19 who remained unvaccinated had a cumulative incidence of COVID-19 that was significantly lower than that of those without prior COVID-19 who were vaccinated. Of the 4284 COVID-19 infections during the study period, 3463 (80.8%) occurred among those without prior COVID-19 who remained unvaccinated, and 800 (18.7%) occurred among those without prior COVID-19 who were vaccinated. Thirteen infections (0.3%) occurred among those with prior COVID-19 who remained unvaccinated, and 8 (0.2%) occurred among those with prior COVID-19 who received the vaccine. Expressed differently, 808 infections (18.9%) occurred among the vaccinated (incidence rate 105.37 per 1000 days at risk) compared to 3476 (81.1%) among the unvaccinated (incidence rate 492.92 per 1000 days at risk) for an incidence rate ratio of 0.21 for vaccination versus no vaccination; whereas 21 infections (0.5%) occurred among those with prior COVID-19 (incidence rate 15.19 per 1000 days at risk) compared to 4263 (99.5%) among those without prior COVID-19 (incidence rate 319.62 per 1000 days at risk), for an incidence rate ratio of 0.048 for prior COVID-19 versus no prior COVID-19.

**Figure 3.**
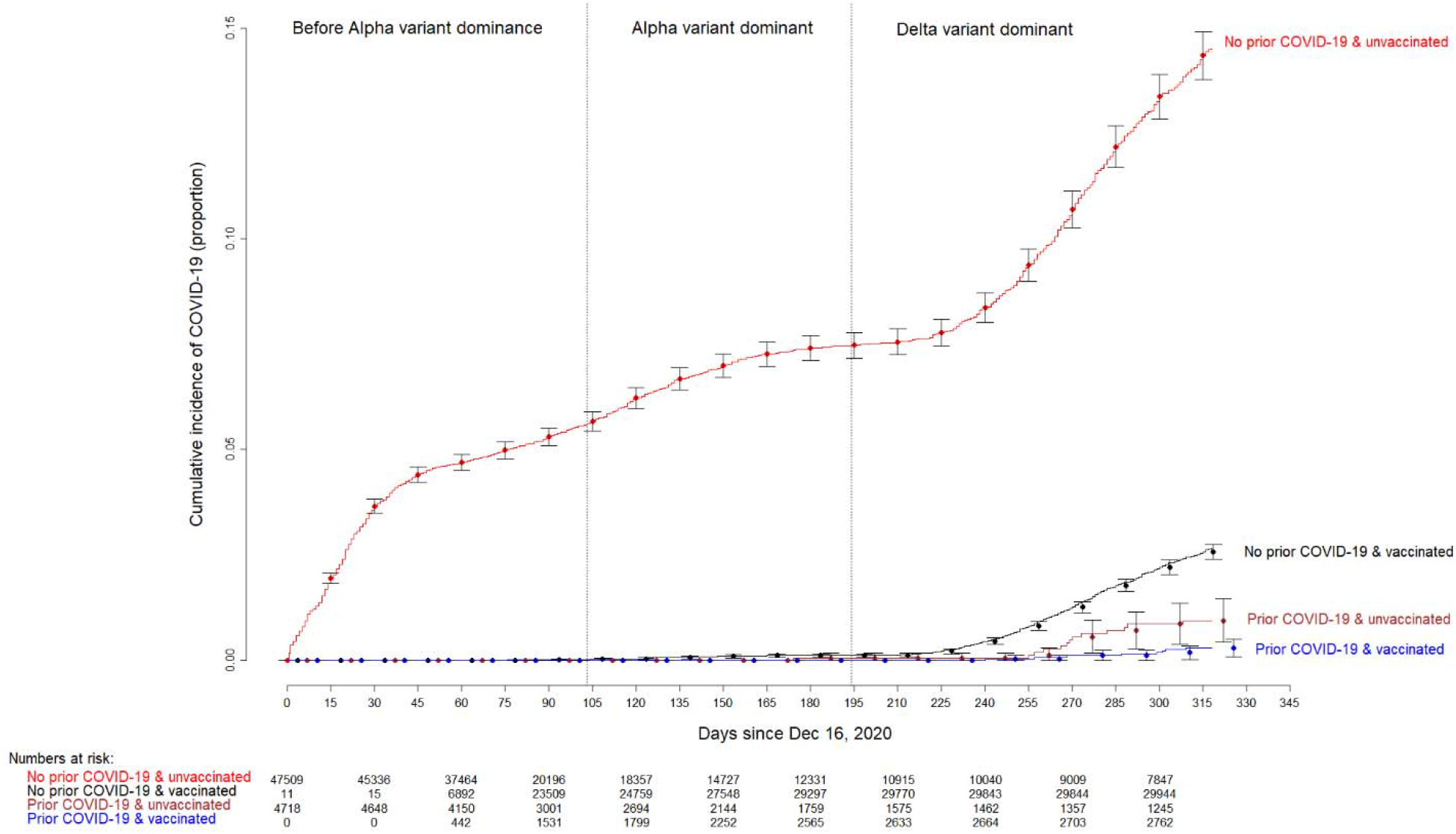
Simon-Makuch plot showing the cumulative incidence of COVID-19 among subjects with and without prior COVID-19, who did and did not receive the vaccine. Day zero was Dec 16, 2020, the day vaccination became available at our institution. Point estimates and 95% confidence intervals are jittered along the x-axis to improve visibility. Eleven subjects who had received two doses of the vaccine before the study start date were presumed to have been vaccinated earlier as participants in clinical trials. A few subjects who received their first dose in the first week of the vaccination campaign received their second dose three weeks later, and were thus considered vaccinated earlier than 42 days. The pandemic phases are identified according to which variant accounted for more than 50% of the strains in HHS region 5 (comprising of Illinois, Indiana, Michigan, Minnesota, Ohio, and Wisconsin) on weekly genomic surveillance conducted by the CDC (https://covid.cdc.gov/covid-data-tracker/#variant-proportions).

Figure 4 shows the cumulative incidence of symptomatic COVID-19 across the various groups, and figure 5 shows the cumulative incidence of hospitalizations for COVID-19. Findings for both symptomatic COVID-19 and hospitalizations for COVID-19 are similar to that of COVID-19 infection overall, in that the group at highest risk was those without prior COVID-19 who remained unvaccinated. Only 8 (0.26%) of the 3092 symptomatic COVID-19 infections occurred among those with prior COVID-19. Not one of the 107 hospitalizations for COVID-19 was among persons with prior COVID-19.

**Figure 4.**
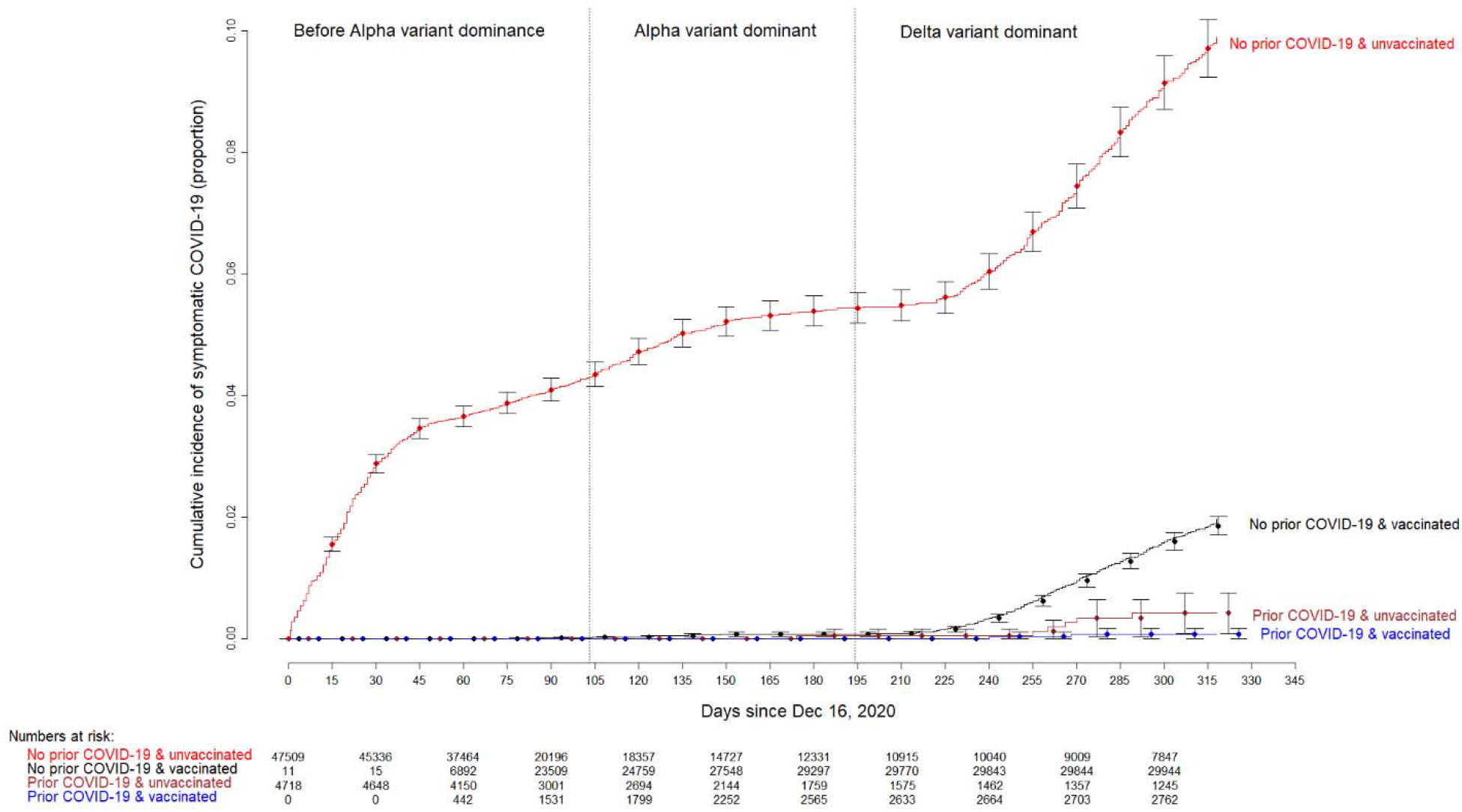
Simon-Makuch plot showing the cumulative incidence of symptomatic COVID-19 among subjects with and without prior COVID-19, who did and did not receive the vaccine. Day zero was Dec 16, 2020, the day vaccination became available at our institution. Point estimates and 95% confidence intervals are jittered along the x-axis to improve visibility. Eleven subjects who had received two doses of the vaccine before the study start date were presumed to have been vaccinated earlier as participants in clinical trials. A few subjects who received their first dose in the first week of the vaccination campaign received their second dose three weeks later, and were thus considered vaccinated earlier than 42 days. The pandemic phases are identified according to which variant accounted for more than 50% of the strains in HHS region 5 (comprising of Illinois, Indiana, Michigan, Minnesota, Ohio, and Wisconsin) on weekly genomic surveillance conducted by the CDC (https://covid.cdc.gov/covid-data-tracker/#variant-proportions).

**Figure 5.**
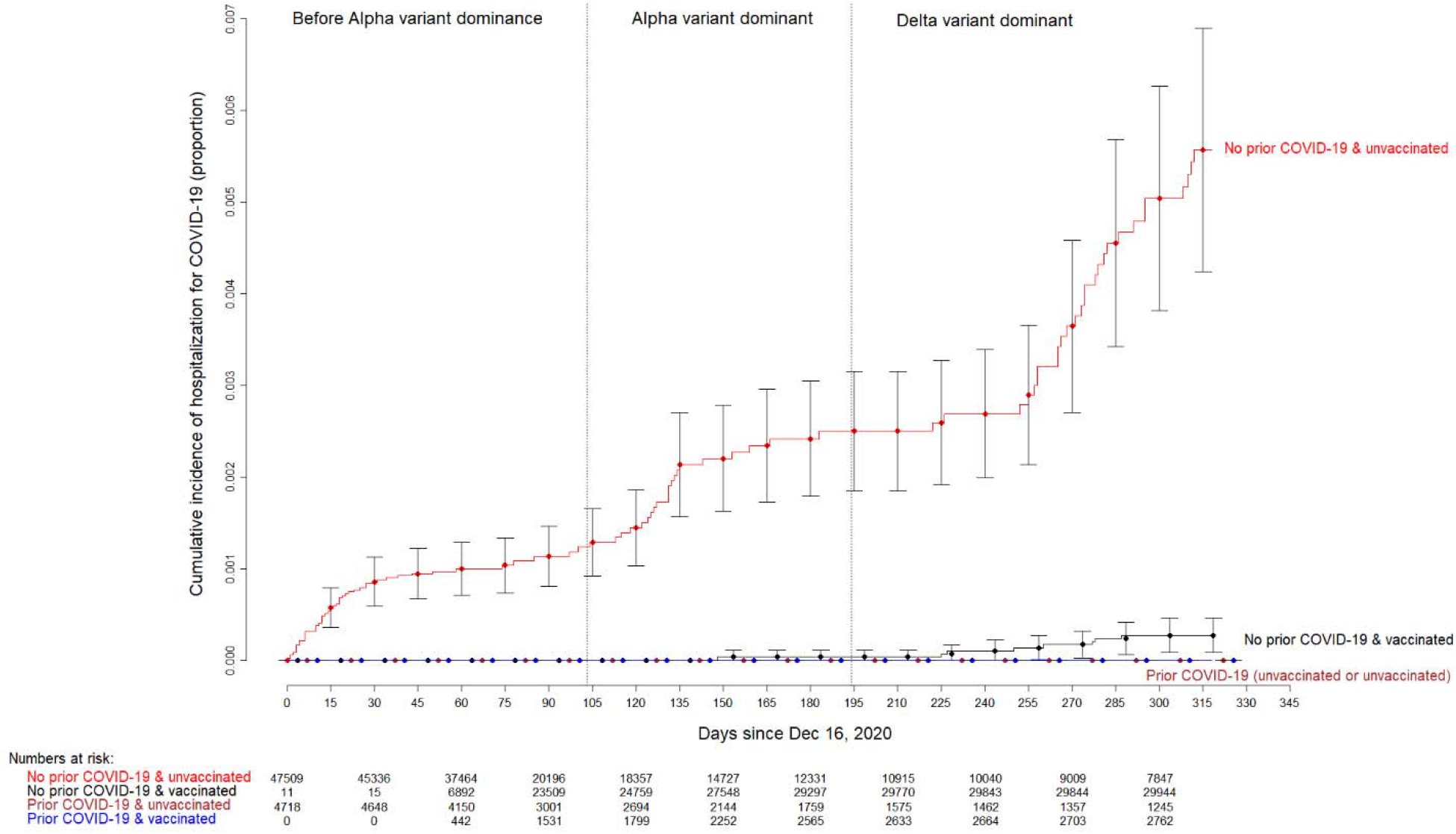
Simon-Makuch plot showing the cumulative incidence of COVID-19 requiring hospitalization among subjects with and without prior COVID-19, who did and did not receive the vaccine. Day zero was Dec 16, 2020, the day vaccination became available at our institution. Point estimates and 95% confidence intervals are jittered along the x-axis to improve visibility. Eleven subjects who had received two doses of the vaccine before the study start date were presumed to have been vaccinated earlier as participants in clinical trials. A few subjects who received their first dose in the first week of the vaccination campaign received their second dose three weeks later, and were thus considered vaccinated earlier than 42 days. The pandemic phases are identified according to which variant accounted for more than 50% of the strains in HHS region 5 (comprising of Illinois, Indiana, Michigan, Minnesota, Ohio, and Wisconsin) on weekly genomic surveillance conducted by the CDC (https://covid.cdc.gov/covid-data-tracker/#variant-proportions).

### Association of prior COVID-19 and vaccination with occurrence of COVID-19

In a Cox proportional hazards regression model, both prior COVID-19 and vaccination were separately associated with significantly lower risk of COVID-19. Increasing age and male gender were also associated with significantly lower risk of COVID-19, and having a patient-facing job was associated with a significantly higher risk of COVID-19.

In a multivariable model, vaccination was associated with a significantly lower risk of COVID-19 among those without prior COVID-19 (HR 0.24, 95% CI 0.22 – 0.26), but this study was not able to find a lower risk of COVID-19 with vaccination among those with prior COVID-19 (HR 0.86, 95% CI 0.33 – 2.29). It is possible that the small numbers of events precluded finding a small relative risk reduction with vaccination among those with prior COVID-19. Given the extremely low reinfection rate among those with prior COVID-19 in the absence of vaccination, even if there is a small relative risk reduction from vaccination, the absolute risk reduction would be very small indeed. Univariable and multivariable associations with time to COVID-19 are shown in table 2.

**Table 2.**
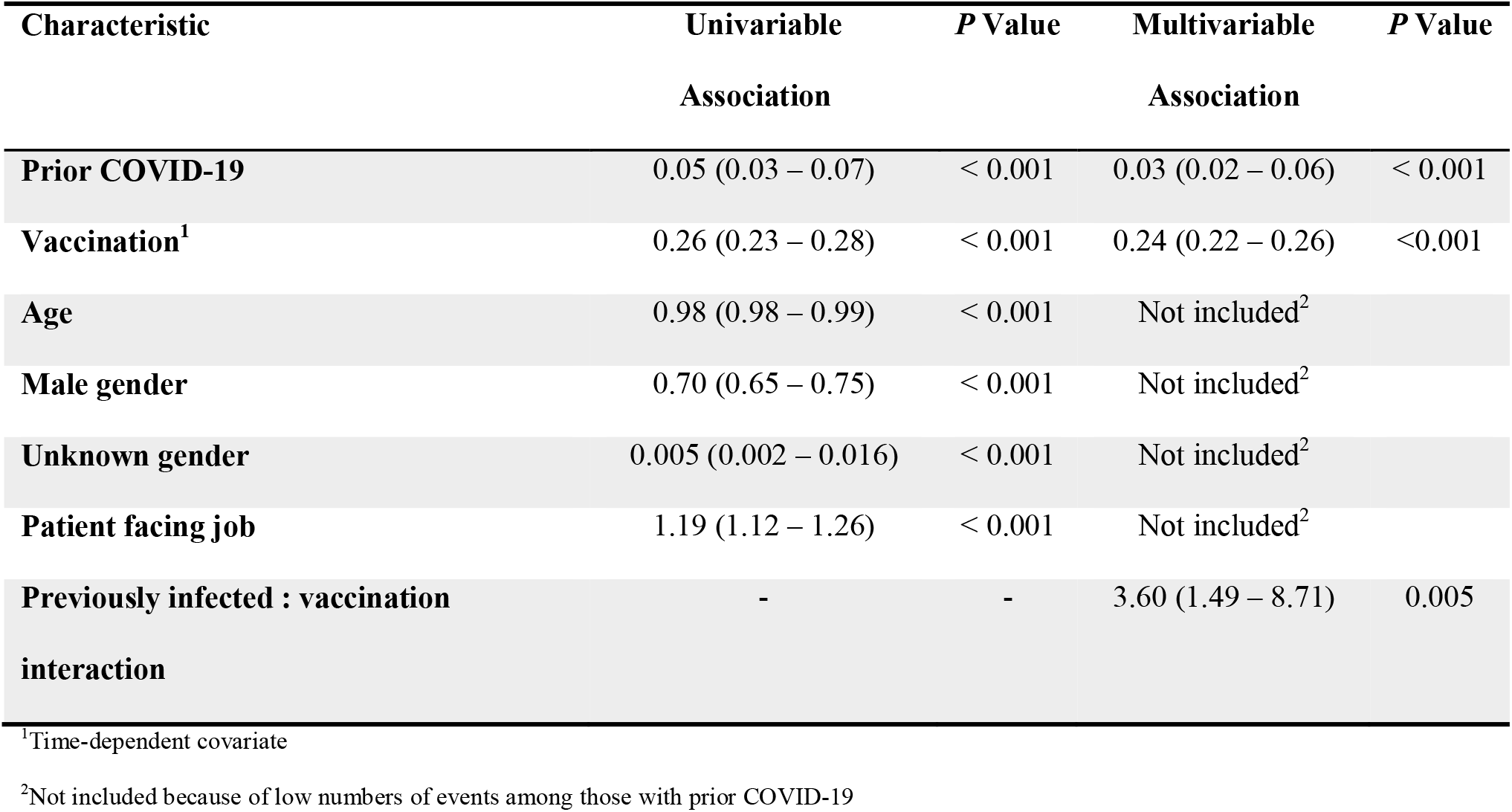
Associations with time to COVID-19.

| Characteristic | Univariable<br>Association | *P* Value | Multivariable<br>Association | *P* Value |
| --- | --- | --- | --- | --- |
| **Prior COVID-19** | 0.05 (0.03 – 0.07) | < 0.001 | 0.03 (0.02 – 0.06) | < 0.001 |
| **Vaccination<sup>1</sup>** | 0.26 (0.23 – 0.28) | < 0.001 | 0.24 (0.22 – 0.26) | <0.001 |
| **Age** | 0.98 (0.98 – 0.99) | < 0.001 | Not included<sup>2</sup> |  |
| **Male gender** | 0.70 (0.65 – 0.75) | < 0.001 | Not included<sup>2</sup> |  |
| **Unknown gender** | 0.005 (0.002 – 0.016) | < 0.001 | Not included<sup>2</sup> |  |
| **Patient facing job** | 1.19 (1.12 – 1.26) | < 0.001 | Not included<sup>2</sup> |  |
| **Previously infected : vaccination**<br>**interaction** | - | - | 3.60 (1.49 – 8.71) | 0.005 |

<sup>1</sup>Time-dependent covariate
<sup>2</sup>Not included because of low numbers of events among those with prior COVID-19

### Duration of protection

This study was not specifically designed to determine the duration of protection afforded by natural infection or vaccination. For those with prior COVID-19 the median duration since prior infection was 33 days (IQR 16 – 119 days), and there were almost no SARS-CoV-2 infections over the following 8 months while infections occurred among those without prior COVID-19. This suggests that prior COVID-19 almost certainly provides protection against reinfection for 9 months or longer. Even though there was a small risk of reinfection among those with prior COVID-19 during the Delta surge, this study was unable to find that vaccination reduced that risk as far out as almost a year since prior infection. The lack of protection by vaccination among those with prior COVID-19 suggests that the increase in infections after 9 months may be because of inherently lower protection of prior infection or vaccination against the Delta variant rather than waning of immunity. Longer follow-up or studies specifically designed to address this question will be needed to clarify how much longer than one year is the duration of protection provided by prior infection.

## DISCUSSION

This study again shows that both individuals with prior COVID-19 and those who received the vaccine are at much lower risk of getting COVID-19 than those without prior COVID-19 who remain unvaccinated. This holds true for prior infection even when it was of a different variant and the current predominant strain circulating in the community is the Delta variant. This holds true even when the prior infection was a year or longer in the past. Additionally, those with prior COVID-19 who did not receive the vaccine were at lower risk of getting COVID-19 than were previously uninfected persons who received two doses of an mRNA vaccine. These findings suggest that it is not necessary to vaccinate those who have already had COVID-19, at least not for a year after their infection, and probably even longer.

This study’s findings should not come as a surprise. It is actually very much in line with what we know about how the immune system works. Individuals exposed to a pathogen they have encountered before are able to mount an immunologic response to protect themselves against reinfection. Earlier observational studies have indeed found very low rates of reinfection over the following months among survivors of COVID-19 [6–14]. Several studies have compared the risk of reinfection among individuals with prior COVID-19 to that of breakthrough infection among vaccinated individuals. A single study conducted by the CDC concluded that prior vaccination was more protective against future COVID-19 than was prior COVID-19 [15]. This finding has not been replicated elsewhere. Multiple observational studies done in different continents comparing the protective effect of prior infection and vaccination against subsequent COVID-19 have all found that both provided similar levels of protection [16–18]. A large nationwide study in Israel found that unvaccinated individuals with prior COVID-19 actually had significantly lower risks of COVID-19, symptomatic COVID-19, and hospitalization, than vaccinated individuals without prior COVID-19 [19]. Our study adds to these by demonstrating that among those with prior COVID-19, adding a vaccine does not appear to add protection against future COVID-19 in addition to the substantial protection already provided by virtue of the prior infection. Additionally, our study was concordant in its finding with the aforementioned Israeli study [19] in that unvaccinated persons with prior COVID-19 had a lower risk of subsequent COVID-19 than did vaccinated persons without prior COVID-19. A study in a healthcare worker cohort in India, also did not find evidence of additional protection with vaccination among 1449 individuals with prior COVID-19, over a 45-day period during a surge of infection [20], a finding that is consistent with what was observed in our study. While both reinfections in persons with prior COVID-19 and breakthrough infections in vaccine recipients occur, our study, similar to other like studies, demonstrates that the latter is far more common, which is also consistent with what is observed in clinical practice.

The strengths of our study include its large sample size and follow-up of up to 10.5 months, including follow-up of more than 11.5 months since prior infection among more than half of those previously infected, and a follow-up period that included the time when the Delta variant was the predominant circulating strain. Given that this was a study among employees of a health system, and that the health system had policies and procedures in recognition of the critical importance of keeping track of the pandemic among its employees, we had an accurate accounting of who had COVID-19, when they were diagnosed with COVID-19, who received a COVID-19 vaccine, and when they received it. Additionally, the organization continued to emphasize the need to seek testing if suspicious symptoms occurred, even in those who had been vaccinated, through intranet home page messages and organization-wide email reminders.

The study has its limitations. Because we did not have a policy of asymptomatic employee screening, previously infected subjects who remained asymptomatic might have been misclassified as previously uninfected. Any such misclassification, however, would have had the effect of underestimating the protective effect of prior infection compared to no prior infection, thereby adding to the strength of the study. Also, when evaluating outcomes, many asymptomatic infections would have been missed. But there is little reason to suppose that they would have been missed in the various groups at rates disproportionate enough to change the conclusions drawn from the study. Our study included no children and few elderly subjects, as it consisted of active employees working within a healthcare system, and the majority would not have been immunocompromised. Data governance policies in our institution precluded us from obtaining detailed clinical information on employees. While one cannot generalize this study’s findings to assume that prior infection would provide adequate immunity in these groups, there is also no reason to expect a vaccine to provide additional protection in these same groups, beyond the protection afforded by prior infection. The previously infected individuals in our study also had their prior infection at a time when monoclonal antibody treatment was not available. There might be concern that administration of monoclonal antibodies during COVID-19 might interfere with the mounting of a normal immunologic response to the infection. Given this possibility, it may not be wise to extrapolate this study’s findings to persons who received monoclonal antibody treatment for their prior COVID-19 infection.

A single study conducted by the CDC concluded that vaccination protects against subsequent COVID-19 among persons with prior infection [21]. This case-control study was biased in that protection from subsequent COVID-19 could be easily explained by better adherence to masking and social distancing recommendations among those who chose to receive the vaccine, which was extremely likely in the population that was studied, rather than vaccination. Other studies whose study designs would allow evaluation of benefit of vaccination in previously infected individuals did not find such a benefit [1,20].

The findings from this study should not be misinterpreted to claim that previously infected individuals will never need a vaccine. The duration of effective protection from prior COVID-19 has yet to be defined. Median freedom from reinfection (time from initial infection until end of follow-up) in this study, for those previously infected, was almost a year. A very low cumulative incidence of COVID-19 reinfection up to almost a year has also been demonstrated elsewhere [14]. A practical and useful message would be to consider both prior COVID-19 and vaccination to be adequately protective against future infection, and that people who have had COVID-19 confirmed by a reliable laboratory test do not need the vaccine for at least a year after they were diagnosed with the infection. There are no studies at this time that would suggest these individuals need a vaccine after a year. Additional studies will be needed to determine if such individuals need to be vaccinated, and when.

Our study’s findings have important implications. It is clear that the group at highest risk of getting COVID-19 is that of individuals who never had COVID-19 and remain unvaccinated. Those who had COVID-19 or received the vaccine are at much lower risk. In light of this study’s findings, thinking of the population as “vaccinated” and “unvaccinated” is a less accurate way to classify risk of getting COVID-19 than classifying into “protected” (anyone who’s either had COVID-19 or has been vaccinated) and “vulnerable” (anyone who has neither had COVID-19 nor received the vaccine). An acknowledgment of what has become increasingly obvious, that people who have had COVID-19 are protected from future COVID-19, should raise question about the wisdom of insisting that all previously infected persons should get the vaccine.

Many of the resource-poor countries have weathered almost two years of the pandemic without the benefit of vaccines. A large proportion of the population in some of these countries might have already had the infection. These countries should obtain estimates of the proportion of their populations that have already had COVID-19 to determine the proportions of their populations that are vulnerable versus protected, and thereby appropriately prioritize acquisition of COVID-19 vaccines in relation to their other pressing healthcare needs, rather than embark on an expensive proposition to unnecessarily vaccinate their entire populations with the COVID-19 vaccine.

In conclusion, both previously infected individuals and those who have been vaccinated are substantially protected against COVID-19 infection. Previously healthy adults in the working age population who have already had COVID-19 are unlikely to benefit from COVID-19 vaccination, at least for a year after their infection. We do not know if these findings can be extrapolated to the elderly, to children, and to the immunocompromised.

## Data Availability

Add data produced in the present study are available upon reasonable request to the authors.

## Author contributions

NKS: Conceptualization, Methodology, Validation, Investigation, Data curation, Software, Formal analysis, Visualization, Writing-Original draft preparation, Writing-Reviewing and Editing, Supervision, Project administration.

ASN: Methodology, Formal analysis, Visualization, Validation, Writing-Reviewing and Editing.

PCB: Resources, Investigation, Validation, Writing-Reviewing and Editing.

PT: Resources, Writing-Reviewing and Editing.

SMG: Project administration, Resources, Writing-Reviewing and Editing.

